# AIR-CONDUCTION AND BONE-CONDUCTION REFERENCE THRESHOLD LEVELS – A MULTICENTER STUDY

**DOI:** 10.1101/2024.08.01.24311230

**Authors:** Robert H. Margolis, Victoria Sanchez, Lisa L. Hunter, Aparna Rao, Suzannah Boyle, Lina Motlagh Zadeh, Amelia N. Wong

**Affiliations:** Arizona State University College of Health Solutions, Tempe, AZ; Audiology Incorporated, Arden Hills, MN; University of South Florida Morsani College of Medicine, Tampa, FL; Cincinnati Childrens Hospital Medical Center, Cincinnati, OH

## Abstract

Air--conduction (AC) and bone-conduction (BC) thresholds were measured to evaluate standard reference thresholds and recommend revisions to audiometer standards. AC and BC thresholds were measured from listeners with normal hearing (NH) and sensorineural hearing loss (SNHL) at three sites. NH participants (n = 53) were selected based on age (18 – 25 years), normal AC thresholds, tympanometry, otoscopy, and absence of otologic disease. SNHL participants (n = 49) were selected based on AC thresholds, tympanometry, otoscopy, and absence of otologic disease. AC thresholds obtained from NH listeners averaged 3.7 dB HL. Air bone gaps (ABGs) occurred in NH and SNHL listeners above 2000 Hz and SNHL listeners at 250 Hz. Corrections to standard RETSPLs are recommended. ABGs in listeners without conductive pathology result from incorrect reference threshold levels for frequencies above 2000 Hz. False air-bone gaps increase with hearing-loss magnitude, probably due to effects of ambient and internal noise for low-level bone-conduction stimuli. False ABGs place patients at risk for unnecessary medical and surgical intervention. Reference threshold levels should achieve two objectives: 1) AC thresholds from young NH listeners should average 0 dB HL; 2) ABGs from listeners with normal middle-ear function should average 0 dB.

## I. INTRODUCTION

Beginning in 2010, a series of articles documented the occurrence of air-bone gaps in listeners with normal hearing and listeners with sensorineural hearing loss (SNHL). In a study performed at Cambridge (U.K.) University, listeners with SNHL exhibited air-bone gaps at 4000 Hz averaging 19.3 dB when tested with AMTAS (Automated Method for Testing Auditory Sensitivity) and 13.2 dB when tested with conventional manual pure-tone audiometry (Margolis et al. 2010). The finding was replicated in follow-up studies using both manual and automated audiometry performed in the U.S. (Margolis et al., 2013), Germany (Fröhlich et al. 2018), India (Vijayasarathy and Shetty 2023), and Australia (Eikelboom and Swanepoel, unpublished, data presented in Margolis et al., 2013). These false air-bone gaps result from incorrect reference equivalent threshold force levels (RETFLs) for high-frequency (>2000 Hz) stimuli in the International and American audiometer standards (ISO 389-3-2016; ANSI S3.6-2018). The history and sources of the error were reviewed by Margolis et al. (2019).

False air-bone gaps are potentially dangerous to patients. Vijayasarathy and Shetty (2003) pointed out that incorrect diagnosis of conductive pathology resulting from false air-bone gaps can lead to “unnecessary surgical and/or medical intervention” (p. 57).

The magnitude of false air-bone gaps increases with the degree of hearing loss (measured by the air-conduction threshold at the test frequency). We hypothesize that the reason for the dependence on hearing-loss magnitude is that bone-conduction thresholds for normal-hearing listeners are elevated by ambient and physiological noise (Sivian and White, 1933; Carhart, 1950; Roach and Carhart, 1956). Bone-conduction stimuli presented to hearing-impaired listeners are necessarily presented at higher levels. As a result the stimuli are well above the noise floor and unaffected by it. This results in the dependence of the false 4000-Hz air-bone gap on hearing sensitivity reported by Margolis et al. (2013), Margolis et al. (2019), Wasserman (2018 summarized in Margolis et al, 2019), Vijayasarathy and Shetty (2023), and the current study.

ISO 389-9-2009 specifies preferred test methods for studies that seek to determine reference equivalent threshold levels for incorporation into the audiometer standards. That standard states that the subjects shall be “otologically normal”. Hearing loss is a symptom of ear disease. Therefore listeners with hearing loss are not otologically normal and have been excluded from being participants in studies intended to derive reference threshold levels. In this report the recommended reference equivalent threshold force levels (RETFL) for bone-conduction stimuli are based on threshold data from listeners with SNHL. In their seminal article on normal auditory sensitivity Sivian and White (1933) explained that thermal noise (pressure variations produced by Brownian movement of air molecules) could impose a limit on auditory sensitivity. Carhart (1950) pointed out that bone-conduction thresholds obtained from normal listeners are potentially contaminated by ambient and internal noise, even in sound isolating test exclosures. In this study the procedures specified in ISO 389-9 were carefully followed with listeners with both normal hearing and SNHL.

The goals of this study are a) provide additional evidence of false air-bone gaps that occur when audiometers are calibrated to standard RETFLs, b) provide recommended bone-conduction reference threshold levels that will eliminate false air-bone gaps, and c) provide recommended air-conduction reference equivalent threshold sound pressure levels (RETSPL) for the most commonly used circumaural earphone (Radioear DD450). Thresholds were obtained at octave and interoctave audiometric test frequencies specified in ISO 389-3:2016 (Table 1) and ANSI S3.21-2004 (par. 6.3). Correctly-defined RETSPLs and RETFLs should result in average air-conduction thresholds of 0 dB HL in young normal-hearing listeners and average air-bone gaps of 0 dB at all frequencies for listeners with normal middle-ear function.

**Table I.** Threshold measurement methods and bone vibrator placement at the three test sites (ASU – Arizona State University, CCHMC – Cincinnati Children’s Hospital Medical Center, USF – University of South Florida. AMTAS = Automated Method for Testing Auditory Sensitivity.

|  | **Air**<br>**Conduction**<br>**Method** | **Bone**<br>**Conduction**<br>**Placement** | **Bone**<br>**Conduction**<br>**Method** |
| --- | --- | --- | --- |
| **ASU** | AMTAS | Mastoid | Manual |
| **CCHMC** | Manual | Forehead | Manual |
| **USF** | AMTAS | Forehead | Manual |

There is an increasing use of automated threshold testing in clinical audiology. A growing number of studies have demonstrated that properly validated automated threshold testing procedures produce results that are equivalent to results of manual testing performed by expert testers (Eikelboom et al., 2013; Mahomed et al., 2013; Margolis et al., 2010; Margolis & Moore, 2011). Automated testing is more efficient when bone-conduction testing is conducted with the bone vibrator placed on the forehead so that the transducer does not need to be repositioned during the evaluation. Calibration levels for forehead bone-conduction testing have been in the standards for many decades and produce equivalent results when the standard conversion values are applied. This investigation employed manual and automated threshold methods and forehead and mastoid bone conduction testing to reflect the various, well-accepted methods used in clinical testing.

## II. METHOD

### A. Data Collection Sites

Data were collected at three sites. Institutional review board approval was obtained at each site: Arizona State University (ASU; IRB approval STUDY00014515), Cincinnati Children’s Hospital Medical Center (CCHMC; IRB approval 2009-0855), and University of South Florida (USF; IRB approval STUDY000265).

### B. Participants

An oral case history obtained information related to ear disease, noise exposure, and family history following the questionnaire in ISO 389-9 (Annex A). Participants in the normal-hearing group were otologically normal as defined in ISO 389-9 (par. 4.1.3) and Møller (1996). Specific criteria for normal-hearing participants were a) age 18-25 years; b) air-conduction thresholds ≤ 20 dB HL at octave and interoctave frequencies 250 – 8000 Hz; c) normal tympanograms (tympanometric peak pressure ≥50 daPa; static admittance at 226 Hz ≥ 0.4 mmho; tympanometric width at 226 Hz < 200 daPa); d) otoscopy – no ear-canal occlusion or other abnormality; e) case history – no prolonged or frequent exposure to high level noise; no prolonged exposure to ototoxic medications; no family history of genetic hearing loss; no known history of otologic disease that may cause permanent threshold shift. In addition to criteria c, d and e, additional criteria for participants with SNHL were a) age 18-80 years; air-conduction threshold ≥ 35 dB at one or more frequencies above 1000 Hz; b) no known history of otologic disease that may cause permanent threshold shift; c) no external or middle-ear infections in the past five years. A total of 53 participants with normal hearing (30 female, 23 male; mean age = 22 yrs; SD = 2.2 yrs) and 49 participants with SNHL (21 female, 28 male; mean age = 62 yrs; SD = 15.7 yrs) were tested.

### C. Procedures

Air-conduction and bone-conduction thresholds were tested at standard octave and interoctave frequencies (250 – 8000 Hz). Air-conduction testing was performed with AMTAS at two sites and by manual audiometry at one site. Bone-conduction thresholds were obtained by manual audiometry at all three sites. Bone-conductor placement was mastoid at one site and forehead at two sites. Table I shows threshold measurement methods and bone-conductor placement utilized at the three sites.

Air-conduction testing was performed with Radioear DD450 circumaural earphones calibrated to Sennheiser HDA 200 RETSPLs in ANSI S3.6-2018 and ISO 389-8-2004. A comparison study demonstrated the equivalence of RETSPLs derived for the two circumaural earphones (Smull et al., 2018). Air-conduction thresholds were tested in both ears but data for one ear (randomly selected) of each subject (equal numbers of right and left ears) were analyzed. Randomization was based on each subject’s study number – e.g. right ear was selected for even numbers; left ear for odd numbers. Bone-conduction thresholds were analyzed for the ear that was analyzed for air conduction. The ear that was analyzed was randomly selected (rather than best ear) because selection of the best ear would produce a biased sample of the population, as demonstrated for audiometric data by Chen et al., 2022.

AMTAS is a computer-controlled threshold-measurement method that utilizes a single-interval, YES-NO, forced-choice paradigm with feedback. Stimuli are presented with a bracketing technique similar to the Hughson-Westlake method (Jerger and Carhart, 1959). Stimulus levels descend in 10-dB steps until a “no” response occurs and then brackets in 5-dB steps. Threshold is defined as the level at which two YES responses each following NO responses occur within a 5-dB range. If the two NO responses occur at different levels the higher of the two levels is defined as threshold. The higher of the two levels was chosen because it produced better agreement with manual audiometry. Manual audiometry was performed with a modified Hughson-Westlake method described in ANSI S3.21-2004.

Bone vibrator placement was mastoid or forehead (Table I). International and American standards provide mastoid – forehead conversion factors (ISO 389-3-2016 Annex C; ANSI S3.6 – 2023 Table 8). Bone-conduction thresholds were tested in one ear with a Radioear B-81 bone vibrator at the same frequencies as air-conduction testing. Bone-conduction testing was performed with the bone vibrator secured by an elastic headband designed to exert the proper force (5.4 N on an average adult head size; ISO-389-3 – par. B.4; ANSI S3.6-2023 – par 9.4.3) on the bone vibrator (Margolis and Margolis, 2022), the test ear uncovered, and the non-test ear covered with the circumaural earphone to deliver masking. A narrow-band masking noise was delivered to the non-test ear at 20 dB above the signal level. All testing was performed in sound-attenuating rooms that meet the requirements for maximum permissible ambient noise levels for air-conduction and bone-conduction audiometry (ISO 8253-1 par. 11; ANSI S3.1 par.4). Margolis et al., (2022) calculated maximum permissible noise levels (MPANLs) for circumaural earphones like those used in the present study. The ambient noise attenuation provided by circumaural earphones provides increased protection against effects of ambient noise.

Bone-conduction stimuli were calibrated with mastoid RETFLs. The forehead-mastoid conversion factors (ANSI S3.6-2018, Table 8, p. 33; ISO 389-3-2016, Table C.1, p. 10) were applied to the USF and CCHMC bone-conduction thresholds that were obtained with forehead placement to convert the values to equivalent mastoid thresholds.

Bias in bone-conduction testing has been demonstrated in manual pure-tone audiometry (Margolis et al., 2016). The bias results from the tester’s knowledge of air-conduction thresholds when testing bone conduction. In this study bias was controlled by several methods. At the ASU and USF sites, bone conduction was tested manually with the tester having no knowledge of air-conduction thresholds. At the CCHMC site where bone conduction was tested with forehead placement, bone conduction was calibrated to mastoid values so that the tester did not know the actual bone-conduction stimulus level and was told not to expect air and bone-conduction thresholds to agree, a method used by Fröhlich et al. (2016). Bone-conduction thresholds were converted to mastoid levels with the conversions presented in the standards.

### D. Calibration

Audiometers at the three sites were calibrated by e3 Diagnostics (Arlington Heights IL) with equipment, methods, and reference threshold levels specified in ANSI S3.6-2018 (Section 9.1-9.4, pp. 25-32) and ISO 389-9 2009 (Section 5.2.3, p. 4). The same equipment was used to calibrate all audiometers. The B&K 4930 Artificial Mastoid and Larson Davis System 824 Sound Level Meter were calibrated by the manufacturers within one year of the audiometer calibration dates. Sound room ambient noise levels were measured at each site to assure compliance with maximum allowable levels.

The Radioear DD450 earphones used at each site were calibrated on an ISO 60318 coupler with a 1/2-inch Bruel and Kjaer condenser microphone (B&K 4180). The sound level meter calibration was checked and adjusted if necessary with the Bruel & Kjaer sound calibrator (B&K 4231) prior to calibration of each audiometer.

The Radioear B-81 bone vibrators were calibrated on the Artificial Mastoid with a coupling load of 5.4 N. The coupling load was measured and adjusted with the calibrated spring balance (B&K UA-0247) provided by the manufacturer. The bone vibrator was positioned on the Artificial Mastoid with the centering tool provided by the manufacturer.

### E. Statistical Analysis

Differences between air-conduction thresholds and air-bone gaps were analyzed separately for the normal-hearing and SNHL groups by analysis of variance with post-hoc analysis to identify test frequencies where differences occurred. Differences between air-bone gaps for the two groups and differences among the three sites were also analyzed. Statistical tests were performed with SPSS ( IBM SPSS Statistics for Macintosh, Version 28.0. Armonk, NY: IBM Corp., Released 2021). Repeated-measures ANOVA was completed with mode (air conduction vs. bone conduction) and frequency (250 Hz, 500 Hz, 750 Hz, 1000 Hz, 1500 Hz, 2000 Hz, 3000 Hz, 4000 Hz, 6000 Hz, 8000 Hz) as within-subject factors and test site as a between-subjects factor. Percentiles (5^th^ and 95^th^) were calculated by the PERCENTILE.INC function of Microsoft Excel (Version 2402 Build 16.0.17328.20124) 64-bit).

## III. RESULTS

### A. Average Thresholds and Air-Bone Gaps

Table II shows means, standard deviations, 5^TH^ percentiles, and 95^th^ percentiles of air-conduction thresholds, bone-conduction thresholds, and air-bone gaps for the two participant groups. For the participants with normal hearing the main effect of frequency was significant (F(9, 42) = 3.7, p <.001).

**Table II.** Means (dB HL), 5^th^ percentiles (dB HL), and 95^th^ percentiles (dB HL) of air-conduction (AC) thresholds, bone-conduction (BC) thresholds, and air-bone gaps (ABG) for normal hearing (NH) and sensorineural hearing loss (SNHL) participants. Standard Deviations (SD) are in dB.

|  |  | FREQUENCY (Hz) |  |  |  |  |  |  |  |  |  |
| --- | --- | --- | --- | --- | --- | --- | --- | --- | --- | --- | --- |
|  |  | 250 | 500 | 750 | 1000 | 1500 | 2000 | 3000 | 4000 | 6000 | 8000 |
| NH | n | 53 | 53 | 52 | 53 | 53 | 53 | 53 | 53 | 53 | 53 |
|  | Mean AC | 4.6 | 3.5 | 4.5 | 3.3 | 3.0 | 2.8 | 5.7 | 3.0 | 1.6 | 5.1 |
|  | SD | 7 | 7 | 6 | 5 | 6 | 6 | 7 | 6 | 6 | 7 |
|  | 5th %ile | -5 | -5 | -5 | -5 | -5 | -5 | -5 | -5 | -5 | -5 |
|  | 95th %ile | 15 | 17 | 15 | 12 | 15 | 10 | 17 | 12 | 10 | 15 |
|  | Mean BC | 2.7 | 4.5 | 5.3 | 5.0 | 4.4 | 7.5 | 0.6 | -6.4 | -4.2 | -7.8 |
|  | SD | 11 | 10 | 9 | 8 | 8 | 9 | 9 | 9 | 6 | 7 |
|  | 5th %ile | 19 | 20 | 18 | 17 | 15 | 21 | 14 | 7 | 5 | 5 |
|  | 95th %ile | -13 | -10 | -9 | -5 | -8 | -10 | -10 | -24 | -11 | -17 |
|  | Mean ABG | 1.9 | -1.0 | -0.6 | -1.7 | -1.4 | -4.7 | 5.1 | 9.4 | 5.8 | 12.9 |
|  | SD | 10 | 8 | 8 | 7 | 9 | 9 | 11 | 9 | 8 | 9 |
|  | 5th %ile | -14 | -15 | -15 | -15 | -15 | -20 | -10 | -4 | -5 | 0 |
|  | 95th %ile | 16 | 9 | 11 | 10 | 10 | 10 | 25 | 24 | 18 | 25 |
| SNHL | n | 49 | 49 | 45 | 49 | 45 | 49 | 46 | 49 | 49 | 48 |
|  | Mean AC | 18.1 | 21.1 | 27.9 | 29.7 | 38.0 | 40.6 | 49.5 | 51.7 | 49.1 | 53.3 |
|  | SD | 17 | 16 | 16 | 16 | 16 | 15 | 14 | 14 | 19 | 24 |
|  | 5th %ile | 50 | 53 | 54 | 58 | 59 | 63 | 74 | 70 | 73 | 88 |
|  | 95th %ile | -5 | 0 | 5 | 5 | 10 | 15 | 30 | 30 | 17 | 10 |
|  | Mean BC | 7.5 | 20.0 | 26.7 | 30.2 | 39.2 | 43.1 | 42.4 | 38.9 | 31.8 | 23.3 |
|  | SD | 14 | 18 | 19 | 19 | 15 | 16 | 14 | 13 | 17 | 21 |
|  | 5th %ile | 33 | 50 | 56 | 60 | 62 | 64 | 65 | 56 | 50 | 54 |
|  | 95th %ile | -10 | -5 | 2 | 0 | 16 | 21 | 21 | 16 | 0 | -18 |
|  | Mean ABG | 9.8 | 0.4 | 0.5 | -0.5 | -0.8 | -2.7 | 6.8 | 13.0 | 15.6 | 31.0 |
|  | SD | 13 | 11 | 10 | 9 | 11 | 10 | 11 | 11 | 12 | 16 |
|  | 5th %ile | -9 | -18 | -10 | -14 | -15 | -15 | -12 | -2 | 0 | 15 |
|  | 95th %ile | 23 | 15 | 15 | 13 | 16 | 15 | 24 | 32 | 31 | 50 |

Pairwise comparisons showed significant differences (p <.05) between means only for 3000 /6000 Hz and 6000/8000 Hz. The main effect of mode (air conduction v. bone conduction) was significant (F[1, 49] = 18, *p* < .01). Post hoc analysis (Tukey post-hoc test with Bonferroni correction) revealed significant differences (P < 0.01) at all frequencies above 1500 Hz indicating significant air-bone gaps at those frequencies. For the SNHL group the main effect of mode was significant (F[1, 37) = 32.2, *p* < .01). Post hoc analysis revealed significant differences (P < 0.05) at 250 Hz and all frequencies above 1500 Hz indicating significant air-bone gaps at those frequencies. Analysis of air-conduction thresholds at the three sites indicated significant differences (F(2,50) = 29, p < .001). Examination of the means (see supplemental digital content) show that the thresholds obtained at the ASU site were higher than the other sites. Air-bone gaps at the three sites did not differ significantly at the 0.05 confidence level (F(2,49) = 3.1, p .054).

Figure 1 (left panel) shows average audiograms with thresholds rounded to the nearest 5 dB for listeners with normal hearing and SNHL. Thresholds are rounded to illustrate the appearance of a typical audiogram obtained with the 5-dB step size specified in the standards on pure tone audiometry (ISO 8253-1; ANSI SS3.21). Average bone-conduction thresholds for normal-hearing listeners were within 5 dB of average air-conduction thresholds for frequencies 250 – 3000 Hz. At 4000 and 8000 Hz larger air-bone gaps were observed.

**Figure 1.**
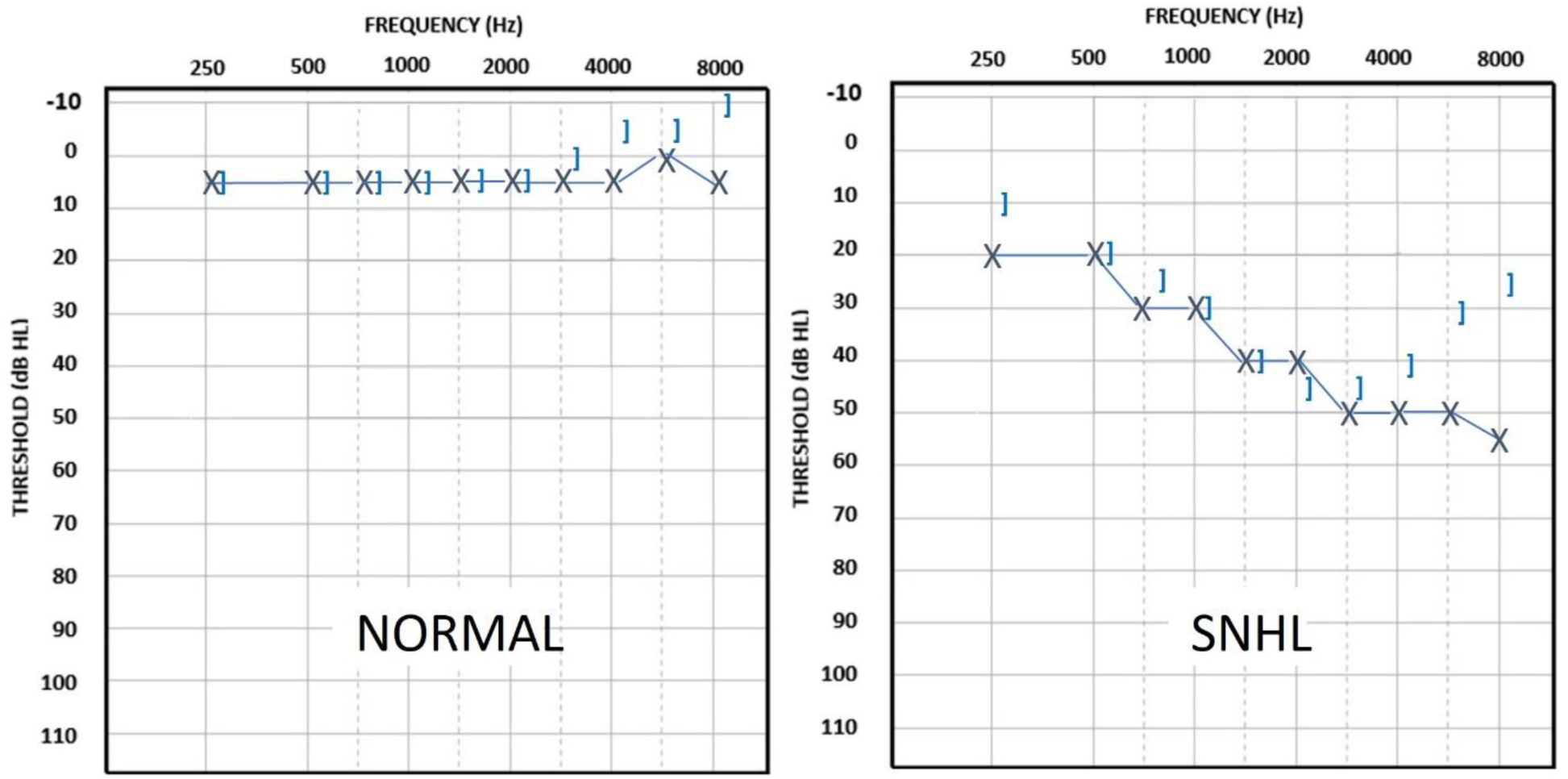
Average audiograms showing thresholds rounded to the nearest 5 dB for the NH and SNHL groups.

Figure 1 (right panel) shows average audiograms with thresholds rounded to the nearest 5 dB for listeners with SNHL. Average bone-conduction thresholds were within 5 dB of average air-conduction thresholds for frequencies 500 – 3000 Hz. At 250 Hz and above 3000 Hz larger air-bone gaps were observed. The air-bone gap tends to increase with frequency above 3000 Hz.

Figure 2 shows unrounded air-bone gaps and standard deviations for the normal and SNHL listeners. The main effect of group (normal v. SNHL) was significant (F[1, 90] = 15.5, *p* < .01). Post hoc analysis revealed significant differences (P < 0.01) at 250, 6000, and 8000 Hz indicating significant differences in air-bone gaps at those frequencies. Standard deviations are smaller for the normal-hearing group than the SNHL group.

**Figure 2.**
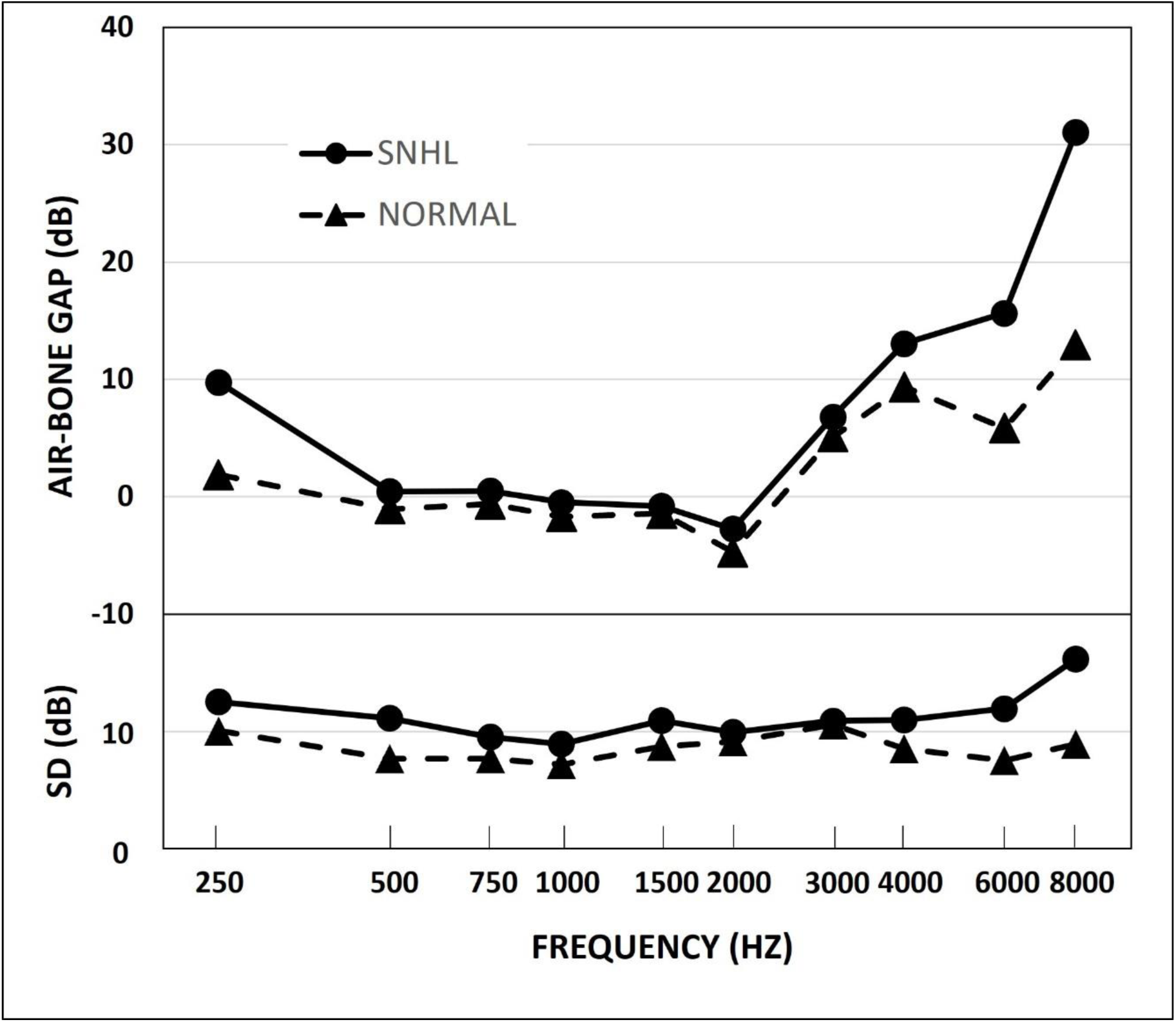
Average (unrounded) Air Bone Gaps and standard deviations (SD) and for the SNHL (circles) and NH (triangles) groups.

The dependence of the air-bone gap on air-conduction hearing sensitivity is shown in Figure 3. Data from the current study are shown along with data from Margolis et al. (2013) and Wasserman (2018 – reported in Margolis et al., 2019). Data from the current study are both groups combined for a total n of 89 cases. Data from the Margolis et al. (2013) study include 637 cases. The best fit linear function determined by linear regression of the weighted average of the three studies (solid line in Figure 3) increases at a rate of 1 dB for each 5 dB increment in air-conduction threshold.

**Figure 3.**
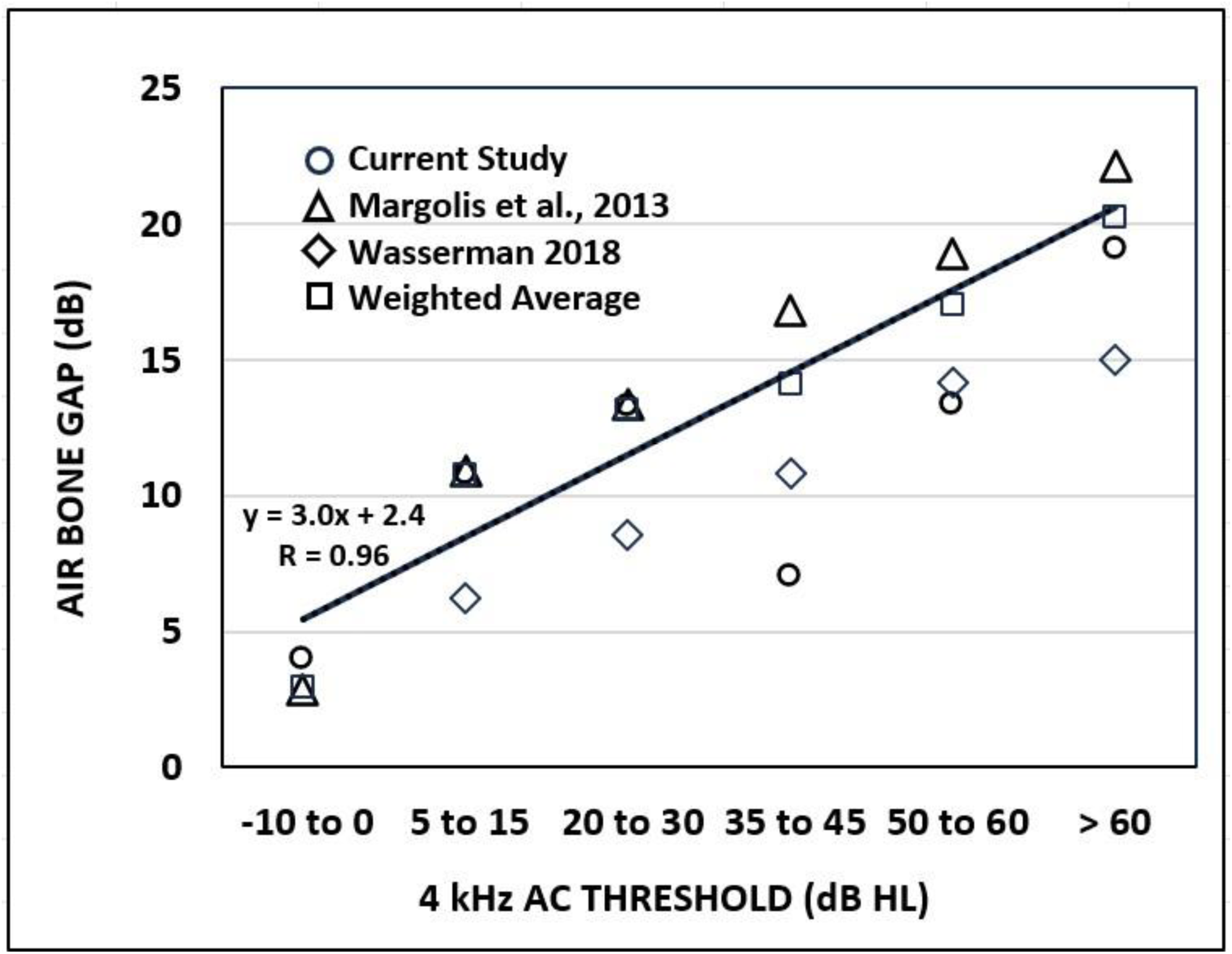
Average Air Bone Gaps plotted against the 4000-Hz AC threshold from three studies. The solid line is the best-fit linear function fit to the weighted average from the three studies.

### B. Reference Threshold Levels

RETSPLs for the DD450 earphone were based on average air-conduction thresholds for the normal-hearing group. ASU air-conduction thresholds for the normal-hearing participants were significantly higher than those from the other sites and from the Smull et al. (2018) data. Therefore they were excluded from the data set used to derive RETSPLs. Weighted average thresholds for the remaining three datasets ( USF, CCHMC, Smull et al. (2018) are shown in Table III. Weighted average thresholds for normal-hearing listeners obtained with the Radioear DD450 earphone calibrated to Sennheiser HDA 200 RETSPLs ranged from −0.6 dB to 4.0 dB with an average across frequencies of 1.2 dB. This is consistent with the comparison study of the two earphones reported by Smull et al. (2018). RETSPLs were calculated by adding the weighted mean thresholds to the HDA200 RETSPLs in the audiometer standards. Corrections, calculated as the weighted mean threshold rounded to the nearest 5 dB, are 0 dB at all frequencies except 8000 Hz where the correction is 5 dB. Corrections are rounded to the nearest 5 dB to conform to the audiogram convention of plotting thresholds in 5 dB steps. The 4-kHz correction can be applied to the threshold value obtained with an audiometer calibrated to the current (ANSI S3.6-2018) standard.

**Table III.** Recommended RETSPLs for the Radioear DD450 earphone. RETSPLs are based on air-conduction thresholds for participants with normal hearing. Corrections are equal to – Weighted Mean Thresholds rounded to the nearest 5 dB.

| Recommended Reference Equivalent Threshold Sound Pressure Levels For Radioear DD450 Earphones |  |  |  |  |  |  |  |  |  |  |
| --- | --- | --- | --- | --- | --- | --- | --- | --- | --- | --- |
| FREQUENCY (Hz) | 250 | 500 | 750 | 1000 | 1500 | 2000 | 3000 | 4000 | 6000 | 8000 |
| n | 43 | 43 | 43 | 43 | 43 | 43 | 43 | 43 | 43 | 43 |
| Weighted Mean Threshold (dB HL) | 0.2 | -0.6 | 1.4 | 0.0 | 1.3 | 1.4 | 2.6 | 1.0 | 0.9 | 4.0 |
| RETSPL (dB re 20 μPa) | 18.2 | 10.4 | 7.4 | 5.5 | 6.8 | 5.9 | 5.1 | 10.5 | 17.9 | 21.5 |
| Correction | 0 | 0 | 0 | 0 | -0 | 0 | 0 | 0 | 0 | -5 |

Table IV shows the recommended RETFLs, adjusted mastoid RETFLs (Mastoid row), and adjusted forehead RETFLs (Forehead row). The values in the correction row are post-hoc corrections that clinicians can employ if their audiometers are calibrated to the current RETFLs. These are the average air-bone gaps for the SNHL group rounded to the nearest 5 dB. These corrections will eliminate false air-bone gaps in most listeners with SNHL.

**Table IV.**
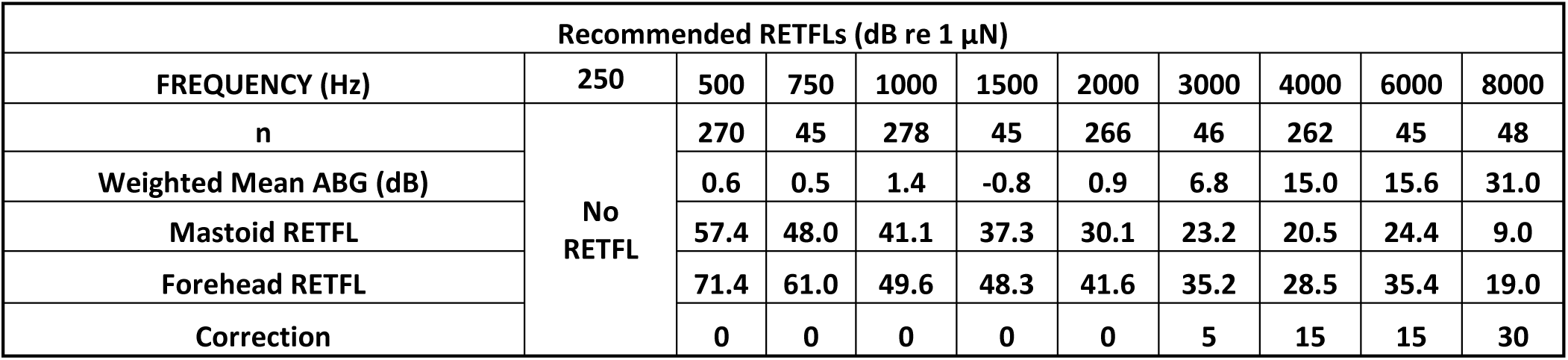
Recommended RETFLs for the Radioear B-81 bone vibrator.

As discussed above, bone-conduction thresholds at 250 Hz are contaminated by two sources: harmonic distortion, and vibrotactile sensitivity. These considerations have led many audiologists to forgo bone-conduction testing at that frequency (Martin et al., 1994; Finan, 2012). While the survey data should not be taken as proof that 250-Hz bone-conduction thresholds do not contribute to the clinical evaluation of patients, they are an indication that audiologists omit testing at that frequency without adverse consequences. Because bone-conduction thresholds at 250 Hz are contaminated by harmonic distortion and vibrotactile sensitivity, we recommend that bone-conduction testing at 250 Hz be omitted from the routine audiometric evaluation. Although the newer B-81 bone vibrator has lower harmonic distortion than the B-71, the distortion at 250 Hz (4%, Radioear B-81 Datasheet) is higher than at other audiometric frequencies and still potentially problematic.

Air-bone gaps at frequencies 500 – 1500 Hz are close to zero for both the normal-hearing group and the SNHL group. No changes in the RETFLs are recommended at those frequencies.

At 2000 Hz small, statistically-significant, negative air-bone gaps were observed for both subject groups (see Table II). The 90% confidence interval for both groups combined was −1.8 to −5.8 dB. Because the confidence interval did not encompass 0 dB, a small correction is recommended.

At frequencies above 2000 Hz air-bone gaps were observed that increased with frequency and are larger for the SNHL group than for the normal group, probably due to the contaminating influence of internal and ambient noise which affects normal-hearing listeners more than listeners with hearing loss. Adjustments to the RETFLs are recommended at those frequencies based on the air-bone gaps observed for the SNHL group.

## IV. DISCUSSION

### A. Sources of False Air-Bone Gaps

The sources of the false air-bone gaps at 250 Hz and at frequencies above 2000 Hz are probably different. At low frequencies, responses to bone-conduction stimuli can be contaminated by harmonic distortion and vibrotactile sensitivity. There are two reasons that responses to 250-Hz tones are more likely to be contaminated by harmonic distortion than responses at higher frequencies. First, there is greater harmonic distortion, and therefore higher distortion product levels, at 250 Hz than at higher frequencies (Dirks and Kamm. 1975; Ginter and Margolis, 2013; Fréden Jansson et al., 2017; Radioear 2023a, 2023b). Second, because the normal bone-conduction threshold is higher at 250 Hz than at higher frequencies, the 250-Hz tones are presented at higher force levels to achieve thresholds so that the harmonics become audible at lower 250-Hz hearing levels. Eichenauer et al., (2014) reported that harmonics of 250-Hz bone-conduction stimuli may become audible for stimuli as low as 20-dB HL with the B71 vibrator and 30-dB HL with the B81 vibrator. In addition, vibrotactile responses to 250-Hz tones presented by bone vibrators may contaminate threshold measurements. Vibrotactile thresholds are significantly lower at 250 Hz compared to higher audiometric frequencies (Boothroyd and Cawkwell 1970; Fréden Jansson et al., 2017; Verillo 1963). Boothroyd and Cawkwell (1970) reported an average vibrotactile threshold at 250 Hz of 37-dB HL in listeners with unilateral hearing loss, suggesting that air-bone gaps in listeners with air-conduction thresholds greater than or equal to 40 dB at 250 Hz may produce responses based on vibrotactile sensitivity to inaudible signals.

Taken together, the influences of harmonic distortion and vibrotactile responses at 250 Hz suggest that bone-conduction thresholds at that frequency may be contaminated and testing is ill-advised. This runs counter to ANSI S3.21-2004-R2023 American National Standard Methods for Manual Pure-Tone Threshold Audiometry which specifies that bone conduction should be tested at octave frequencies from 250 – 4000 Hz. Surveys by Martin et al. (1994) and Finan (2012) reported that 59% and 42% of responding audiologists, respectively, routinely test bone conduction at 250 Hz, suggesting a trend toward decreasing bone-conduction testing at that frequency.

False air-bone gaps at higher frequencies probably have a different source. Early studies of normal bone-conduction sensitivity revealed a simple, elegant relationship between bone-conduction thresholds and frequency. Average thresholds decrease with a slope of −12 dB/octave. (See Corliss et al., 1959 p. 54 shown in Figure 4). The −12 dB/octave slope has a special significance in mechanics. As force decreases at that rate, acceleration remains constant. Corliss et al., (1959) and Whittle (1965) pointed out that the −12 dB/octave slope of the bone-conduction threshold data suggests that bone-conduction sensitivity is determined by the acceleration of structures (like the basilar membrane) responding to sinusoidal displacement. But the frequency dependence of the RETFLs in the standards are not consistent with the findings of Corliss et al. (1959) or its replications (e.g., Studebaker, 1967; Whittle, 1965; Wilber and Goodhill, 1967) that confirmed the −12 dB/octave relationship. This is illustrated in Figure 4 which shows the Corliss data, a line that decreases at −12 dB/octave, and the RETFLs from ANSI S3.6 and ISO 389.3. Where the Corliss average thresholds continue to decrease above 2000 Hz, the RETFLs *increase* (dashed line in Figure 4). The result is a difference between the RETFL and the value predicted by the −12 dB/octave relationship of 14.1 dB at 4000 Hz – very close to the false air-bone gaps at that frequency reported in the several studies cited in this article.

**Figure 4.**
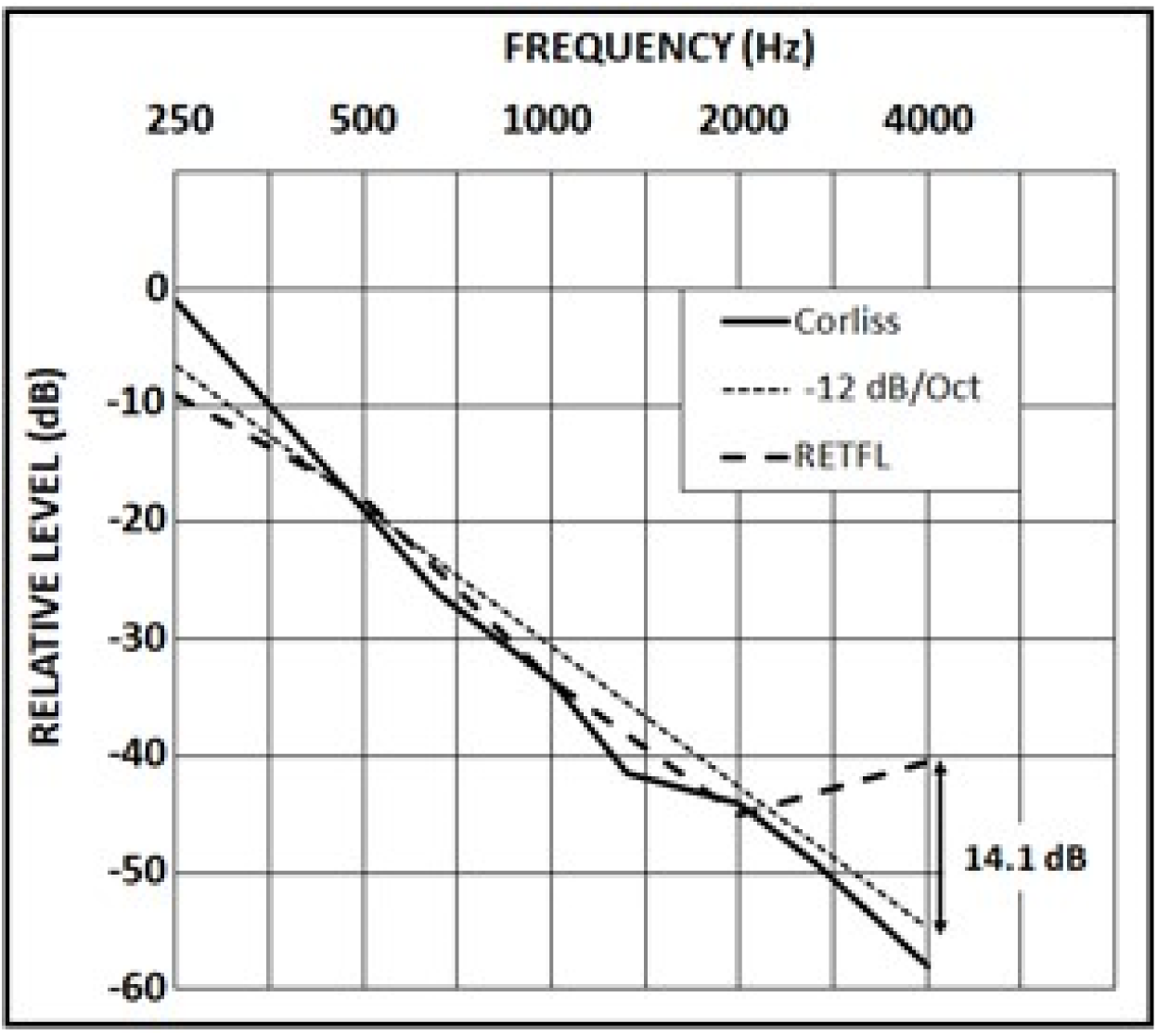
Normal bone-conduction thresholds from Corliss et al. (1959), a best-fit linear function with a slope of −12 dB/octave, and the bone-conduction RETFLs from ANSI S3.6-2018. The difference between the 4000 Hz RETFL and the best fit function is 14.1 dB, close to the false ABGs measured from listeners with SNHL. Values are relative levels (dB) from thresholds expressed in force units.

### B. Dependence of False Air-Bone Gaps on Air-conduction Sensitivity

The foregoing discussion does not explain why the false 4000-Hz air-bone gap increases with the magnitude of the hearing loss in hearing-impaired listeners (Figure 3). We hypothesize that the increase in air-bone gap with increases with hearing loss results from the influence of ambient and physiological noise on bone-conduction thresholds. As the average, normal bone-conduction threshold diminishes at −12 dB/octave with increasing frequency (Figure 4) the stimulus level approaches the noise floor. The noise floor has contributions from ambient acoustic noise, internal physiological noise, and thermal noise. As the signal level approaches the noise floor, the threshold is progressively contaminated, elevating the bone-conduction threshold and decreasing the false air-bone gap. In contrast, the air-conduction thresholds of normal-hearing listeners *increase* above 2000 Hz (see air-conduction RETSPLs in ISO 389-8:2004, Table 1; ANSI S3.6:2018, Table 8; Sivian and White, 1933, Figure 10). With increasing frequency, bone conduction thresholds approach the noise floor while air conduction thresholds move above the noise floor. The decrease in the false air-bone gap due to contamination by internal and external noise is greatest for listeners with low air-conduction thresholds because the bone-conduction thresholds are elevated by the noise floor. For listeners with sensorineural hearing loss the stimuli are above the noise floor and unaffected by it.

Because normal bone-conduction thresholds decrease with increasing frequency as described above and normal air-conduction thresholds increase with increasing frequency above 3000 Hz (see air-conduction RETSPLs in ISO 389-8:2004, Table 1; ANSI S3.6:2018, Table 8) the noise floor at high frequencies affects air and bone-conduction thresholds differently with greater masking for bone-conduction signals than for air-conduction signals. For listeners with SNHL bone-conduction stimulus levels are higher, decreasing the influence of the noise floor. This accounts for the increases in air-bone gaps with increasing hearing loss (Figure 3).

This masking effect can be avoided by selecting participants with SNHL whose thresholds are significantly above, and therefore not masked by, the noise floor as recommended by Carhart (1950) and Roach and Carhart (1956). By basing the RETFLs on results from listeners with SNHL and correcting the high-frequency RETFLs (above 2000 Hz), false air-bone gaps that put these subjects at risk for unnecessary surgery and medical treatment, will be avoided. Although this is a departure from the requirement that participants in studies designed to determine RETFLs be “otologically normal” (ISO 389-9 par. 4.1.3), basing RETFLs on thresholds from participants with sensorineural hearing loss avoids the contamination of thresholds by ambient and physiological noise and results in average air-bone gaps of 0 dB in listeners with normal middle-ear function.

The ISO-389-9 standard that specifies test conditions for studies that are designed to determine RETSPLs and RETFLs requires that the listeners must “otologically normal”. Because hearing loss is a symptom of otologic disease, hearing-impaired listeners have been excluded from studies that determine reference threshold levels. Carhart (1950) and Roach and Carhart (1956) warned against deriving RETFLs from listeners with normal hearing. Carhart (1950) wrote: “False thresholds will also appear in attempts to calibrate the bone unit with normal ears.” Roach and Carhart (1956) described a psychophysical method for calibrating bone conduction with threshold data from listeners with SNHL. The rationale was that ambient noise levels, even in sound-treated test rooms may contaminate bone-conduction threshold measurements in normal-hearing listeners, especially when the test ear is uncovered. Internal noise has a similar effect, especially at high frequencies, as discussed above.

In summary, deriving RETFLs based only on threshold data from normal-hearing listeners is problematic for the following reasons. First, it is common to omit bone-conduction testing for listeners with normal air-conduction hearing. Thus the bone-conduction calibration standards are based on listeners for whom the test is usually not performed. Second, because normal high-frequency bone-conduction thresholds are elevated by the noise floor, as discussed above, the true relationship between air-conduction and bone-conduction sensitivity is obscured in normal-hearing listeners. Third, basing the high-frequency RETFLs on thresholds from normal-hearing listeners will result in false air-bone gaps for listeners with SNHL who are unaffected by the noise floor at high frequencies. For these reasons the recommended RETFLs in this report are based on results from the participants with SNHL. Air-bone gaps at frequencies 500 – 1500 Hz are close to zero for both the normal-hearing group and the SNHL group. No changes in the RETFLs are recommended at those frequencies.

At 2000 Hz small, statistically-significant, negative air-bone gaps were observed for both subject groups (see Table II). The 90% confidence interval for both groups combined was −1.8 to −5.8 dB. Because the confidence interval did not encompass 0 dB, a small correction is recommended.

At frequencies above 2000 Hz air-bone gaps were observed that increased with frequency and are larger for the SNHL group than for the normal group, probably due to the contaminating influence of internal and ambient noise which affects normal-hearing listeners more than listeners with hearing loss. Adjustments to the RETFLs are recommended at those frequencies based on the air-bone gaps observed for the SNHL group.

## V. CONCLUSION

Small differences in air-conduction thresholds obtained with Radioear DD450 earphones and standard RETSPLs for circumaural earphones were observed, mostly within 2 dB. A revised RETSPL is recommended at 8000 Hz only.

Air and bone-conduction thresholds from listeners with normal hearing and SNHL are characterized by false air-bone gaps at 250 Hz and frequencies above 2000 Hz. Contamination of 250-Hz bone-conduction threshold measurements by harmonic distortion and vibrotactile sensitivity result in unreliable threshold measurements. Routine testing of bone conduction at that frequency is not recommended. High-frequency air-bone gaps result from erroneous RETFLs in the standards. Bone-conduction RETFLs for frequencies above 2000 Hz should be based on test results from listeners with SNHL because high-frequency bone-conduction thresholds from normal-hearing listeners are contaminated by effects of ambient and internal noise. Many clinics have adopted bone-conduction correction factors for patients with hearing loss due to the known error in high-frequency RETFLs because the occurrence of false air-bone gaps can result in unnecessary medical and surgical treatment. To avoid these false air-bone gaps revised RETFLs are recommended. The international and American standards should be modified to eliminate false indications of middle-ear disease which put patients at risk.

## AUTHOR DECLARATIONS

The authors have no conflicts of interest to declare. Institutional review board approval was obtained at each test site: Arizona State University (ASU; IRB approval STUDY00014515), Cincinnati Children’s Hospital Medical Center (CCHMC; IRB approval 2009-0855), and University of South Florida (USF; IRB approval STUDY000265). Portions of this work were presented at the annual conference of the American Auditory Society (AAS) in Scottsdale, AZ, March 3, 2023, The American Academy of Audiology (AAA) in Seattle, WA, April 21, 2023, and the 8th International Congress on Bone Conduction Hearing and Related Technologies (OSSEO) in Denver, CO, September 7, 2023.

## Data Availability

All data produced in the present work are contained in the manuscript

The Individual threshold data from this study and further methodological details are available at medRxiv (Margolis et al., 2024, DOI 10.1101/2024.08.01.24311230).

## ACKNOWLEDGEMENTS

We are grateful to Travis McColley and Steve Wood for their assistance with calibration of the audiometers used in this study. Richard Wilson provided a very helpful review of the manuscript.

## SUPPLEMENTAL DIGITAL CONTENT

1. Test Sites, Equipment, Bone Conduction Threshold Corrections Arizona State University Project Coordinator – Aparna Rao, Ph.D. Audiometer – GSI Audiostar Pro Earphones – Radioear DD450 Bone Vibrator – Radioear B81 Bone Conduction Corrections: None Cincinnati Childrens Hospital Medical Center Project Coordinator – Lisa L. Hunter, Ph.D. Audiometer – Interacoustics Equinox Earphones – Radioear DD450 Bone Vibrator – Radioear B81 Bone Conduction Corrections: Forehead thresholds converted to mastoid (see corrections below) University of South Florida Morsani College of Medicine Project Coordinator – Victoria Sanchez, Ph.D. Audiometer – GSI Audiostar Pro Earphones – Radioear DD450 Bone Vibrator – Radioear B81 Bone Conduction Corrections: Forehead thresholds converted to mastoid (see corrections below)
2. Forehead – Mastoid Bone Conduction Corrections From ANSI S3.6 – 2018 Table 8 p. 33 and ISO 389-3-2016, Table C.1, p. 10

3. Individual Threshold Data See Tables S1 and S2

| Frequency (Hz) | 250 | 500 | 750 | 1000 | 1500 | 2000 | 3000 | 4000 | 6000 | 8000 |
| --- | --- | --- | --- | --- | --- | --- | --- | --- | --- | --- |
| Forehead-Mastoid (dB) | 12.0 | 14.0 | 13.0 | 8.5 | 11.0 | 11.5 | 12.0 | 8.0 | 11.0 | 10.5 |

---

**Table S1.**
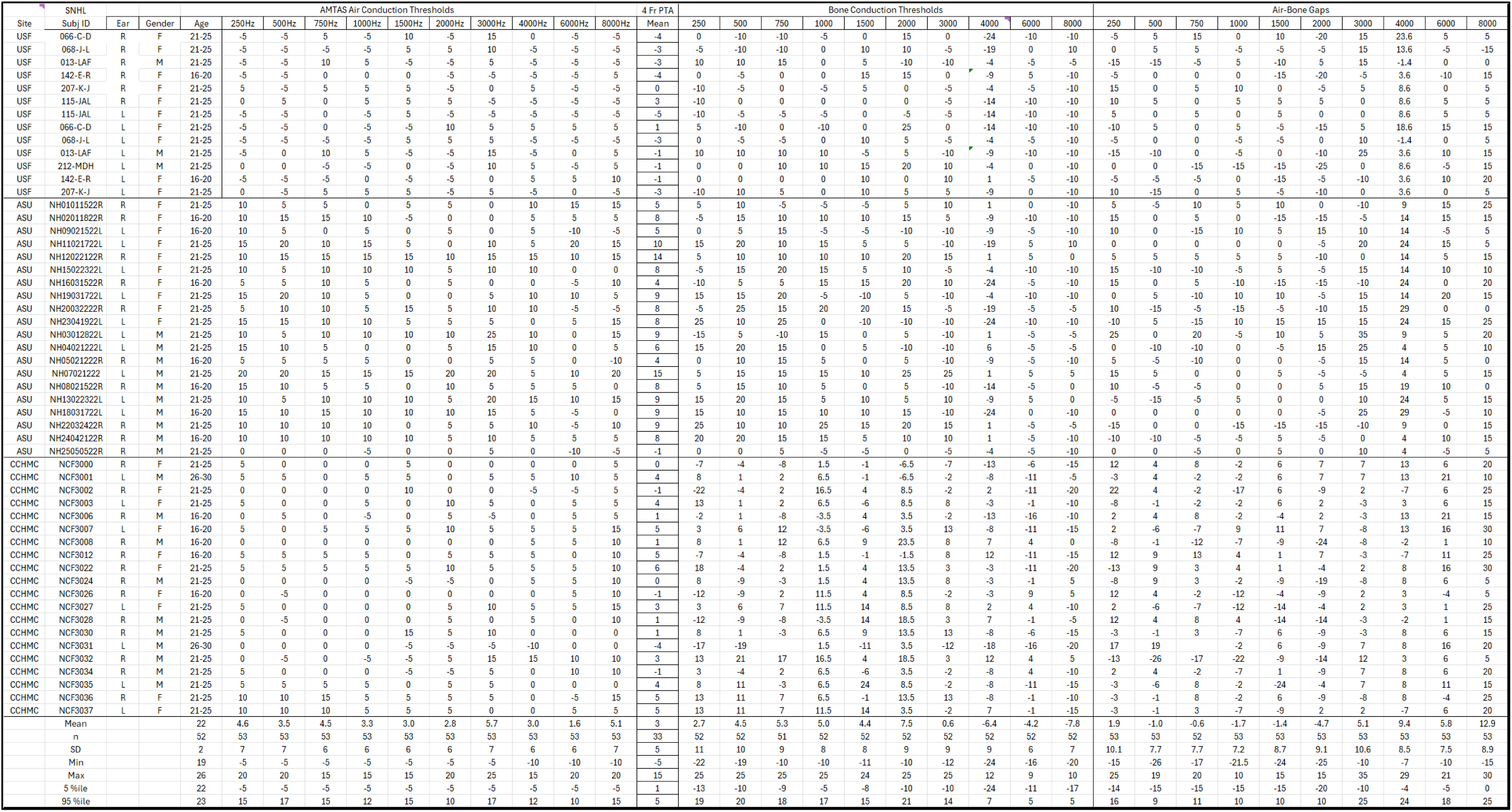
Individual Data for Normal-Hearing Listeners.

**Table S2.**
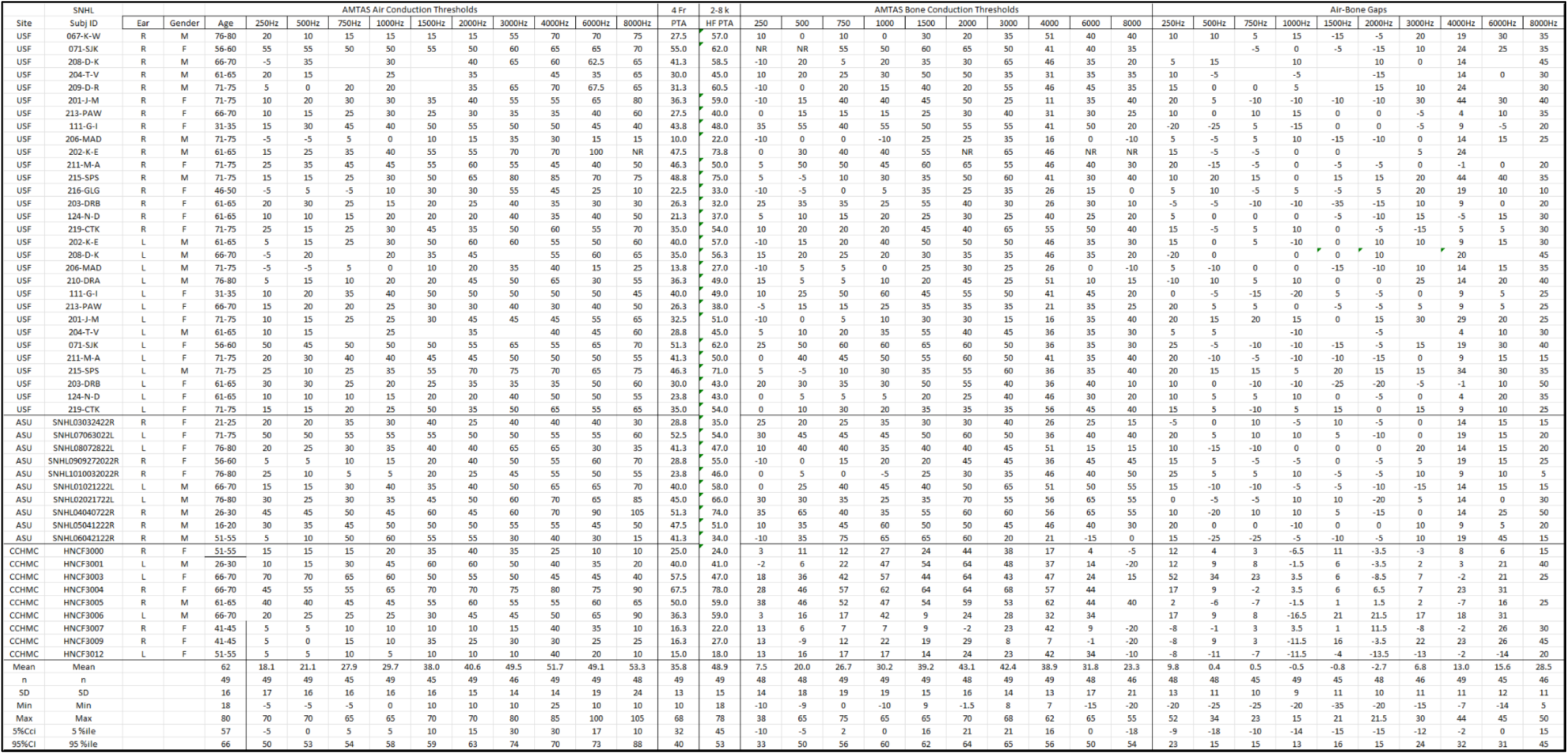
Individual Data for Hearing-Impaired Listeners.

